# Prevalence and associated factors of suicidal ideation among adolescents in rural Bangladesh

**DOI:** 10.1101/2024.05.30.24308223

**Authors:** Rifa Tamanna Mumu, Md Parvez Shaikh, Dipak Kumar Mitra

**Affiliations:** Department of Public Health, School of Health and Life Science, North South University, Bashundhara, Dhaka, Bangladesh; Industrial and Production Engineering, Shahjalal University of Science and Technology, Sylhet, Bangladesh

**Keywords:** Suicidal behavior, Adolescents, Rural Bangladesh, Adolescent mental health, Suicide, Mental health

## Abstract

**Background:** Suicide ranks as the fourth most common cause of death globally, which is more pronounced in lower-middle-income countries. LMICs witness 88% of adolescent suicides yearly, with a 14% prevalence of suicidal ideation. A few studies are available based on the suicidal behavior of young adults residing in rural Bangladesh.

**Objective:** This study seeks to evaluate the prevalence and associated factors of suicidal ideation in 11 to 17-year-old adolescents in a remote subdistrict in Bangladesh.

**Method:** A cross-sectional study was performed in Lohagara, a rural subdistrict in Narail, in southern Bangladesh from April, 15 to May 14, 2024. 350 subjects were recruited for the study, all of whom were school-going adolescents aged 11 to 17 years. The Bengali-translated versions of the Suicidal Behavior-Revised Questionnaire (SBQ-R) and the Depression, Anxiety, and Stress Scale - 21 Items (DASS-21), as well as another structured questionnaire, were used to collect data from participants. Data analysis was done by STATA version 17.

**Result:** The prevalence of current suicidal ideation among adolescents is 21% (95% CI: 17% to 25.8%). Smoking, lack of close friends, an unfavorable family environment, and depression were found to be significantly associated with the development of suicidal ideation in adolescents. Unmarried and single relationship status played a protective role in growing young adults’ suicidal thoughts.

**Conclusion:** The notable prevalence of suicidal ideation in adolescents underscores the need for screening and intervention at an early age to protect them from dreadful consequences. By shedding light on this issue, different health promotional and educational programs, organized in schools and communities may help raise awareness in students, teachers as well as legal guardians. The ultimate goal is to safeguard adolescents from making devastating decisions with adequate counseling, care, and support.

## Introduction

Annually, around 703 individuals are succumbing to suicide globally, as reported by the World Health Organization. Notably, the rate of adolescent suicide is 88% in lower-middle-income countries like Bangladesh, where 90% of the world’s preadults reside. In the year 2019, suicide ranked as the fourth preeminent cause of death among 15 to 29 aged individuals. Statistics unveil a 9.5% worldwide suicide rate in males and 4.18% in females in the 15 to 19 years age bracket in 2009 with a greater susceptibility in elderly adolescents, males in particular (2).

A study conducted with 12 to 17-year-old young adults from 82 lower-middle-income countries revealed a 14% prevalence of suicidal ideation. It was higher in African regions (21%) and lower in Asian regions (8%). Several factors, including female gender, older age, lower socioeconomic status, lack of close friendships, overprotective parenting, peer conflicts, isolation, and victimization, were notably associated with anxiety and suicidal ideation among adolescents (3). Brazil showed a similar trend, where 11 to 15-year-olds manifested a 14.1% prevalence of suicidal ideation, and associated factors included female gender, substance use, conduct disorders, and depressive symptoms (4). On the other hand, a 21.6% prevalence was evaluated including 21.3% in males and 23.5% in females among rural adolescents in Uganda (5).

In the context of Bangladesh, a comparatively reduced suicide rate can be observed in city dwellers than in country people. Adolescents, the elderly, and rural inhabitants are more susceptible to suicide, the adolescent suicide rate is found 20.1% in rural Bangladesh (6). 5% lifetime suicidal ideation is detected among 14 to 19-year-old country adolescents in Bangladesh, revealed in a study conducted in 2013 (7). The prevalence of suicidal ideation among university students was 13.8% which is influenced by gender, academic year, socioeconomic status, depression, family history of suicide, and exposure to traumatic events (8). Among 18 to 28-year-olds 12.8% rate of suicidal ideation rate was determined during the early-COVID-19 period, where the risk factors encompassed previous suicidal attempts, family history, sleep issues, depression, anxiety, and stress (9).

Research carried out with the Global School-based Health Survey data in Bangladesh (2014) visualized an 11.7% prevalence of suicidal behavior in school-going young adults aged 11 to 18 years. Associated factors included loneliness, bullying, a lack of close friendships, anxiety, substance abuse, participating in sexual activity, experiencing a deficit of parental oversight in homework checking, and sparse peer support (10). Although several investigations in Bangladesh have been conducted to explore adolescent suicidal ideation, the concentration has predominantly been on university students, which has created a gap in reckoning the situation among school-going rural adolescents. Hence, this study aims to evaluate the prevalence and associated factors of suicidal ideation in 11 to 17-year-old adolescents in a rural sub-district in Bangladesh. The knowledge of the current rates and associated factors may help stakeholders and policymakers make appropriate decisions and actions to raise awareness to safeguard adolescents’ mental well-being and protect them from making devastating decisions.

## Method

### Study Design

A cross-sectional study was conducted between Apil 15 and May 14, 2024, at Lohagara Government Pilot High School, Lohagara, Narail to assess the prevalence of suicidal ideation and the factors associated with its development among adolescents in Lohagara, a rural subdistrict in Bangladesh.

### Study Participants

The target population was adolescents aged 11 to 17 years in Lohagara and the sample population was the students studying at Lohagara Government Pilot High School, a high school in Lohagara.

### Study Period

The study was conducted from April 15 to May 15, 2024.

### Sample Size

Considering the prevalence of suicidal behavior among adolescents in Bangladesh is 11.7% according to a study conducted using the Global School-based Survey data of 2014 (10), 95% confidence interval, and a 4% margin of error, calculated sample size, 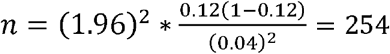

The sampling technique was stratified sampling. Students from different classes were grouped into sections. There were five classes, and each class consisted of three different sections. Every section comprised fifty students in total. Through simple random sampling, one section from each class was selected primarily, and another two sections were chosen from the rest following the same.

### Data Collection Tools

The persistence of suicidal ideation was determined by the Bengali-translated version of the Suicidal Behavior-Revised Questionnaire (SBQ-R), which consists of four questions scoring from 3 to 18. The first question of this questionnaire was used to assess lifetime suicidal ideation.

The Depression, Anxiety, and Stress Scale-21 Items (DASS-21) was used to assess psychological factors like the level of anxiety, depression, and stress. Another structured questionnaire was used to collect data on participants’ sociodemographic, behavioral, socioenvironmental, institutional, and family-environmental factors. Data were collected from respondents translating all questionnaires into the Bengali language. The overall questionnaire was tested on some target population rather than the study participants and the necessary changes were made before data collection.

### Data Management & Analysis

The data for the study was analyzed by STATA version 17. An Independent sample t-test was performed to compare the mean between the two groups and Pearson’s chi-square test was performed to find out the possible association of sociodemographic, behavioral, psychological, family-environmental, institutional, and socio-environmental factors with adolescent suicidal thoughts. To adjust the confounding factors, a multivariate analysis using multinomial logistic regression was performed. Adjusted and unadjusted odds ratios and their 95% CI were used as indicators strength of the association.

### Ethical Considerations

Ethical permission was taken from the Scientific Review Committee(SRC) and Institutional Ethics Committee of North South University before data collection. A permission letter was obtained from the school authorities. Informed written consent was taken from the legal guardians of students before data collection. The respondents were assured of the confidentiality of information. They were also informed that this data would be used for study purposes only.

## Result

### Sociodemographic characteristics of adolescents

A total of 350 data was collected from adolescents with 11 to 17 years of age brackets with a mean age of 14 years (95% CI: 13.8-14.2) from a school in Lohagara. The response rate was 97.4%. More than half of the participants (55.7%) were aged between 11 to 14 years and considered younger adolescents and the rest (44.3%) were comparatively older adolescents aged 15 to 17 years. The proportion of male and female apprentices was almost equal; 51.6% (175) were female and 48.4% (164) were male. Considering the perspectives of monthly family income, the number of family members, house ownership, and parents’ occupations, around half of the participants (48.7%) belonged to families of medium socioeconomic status. 23.8% (81), and 27.5% (94) came from families with high and low socioeconomic status respectively. Most of the students were religiously Muslim; 15% (50) belonged to the Hindu community. A small percentage (4%) were engaged in part-time work apart from study. 78.7% (266) of students were living with their parents, whereas 17.5% (59) were with either father or mother, and 3.9% (13) were without parents. Only 8% (27) of the total had step-parents. A greater portion of students (92%) were unmarried and single. Among the rest, 4.5% (15) were carrying on a good relationship, 2.7% (9) were enduring a complex relationship, and 2 (0.6%) students were married **Table *1***.

**Table 1.**
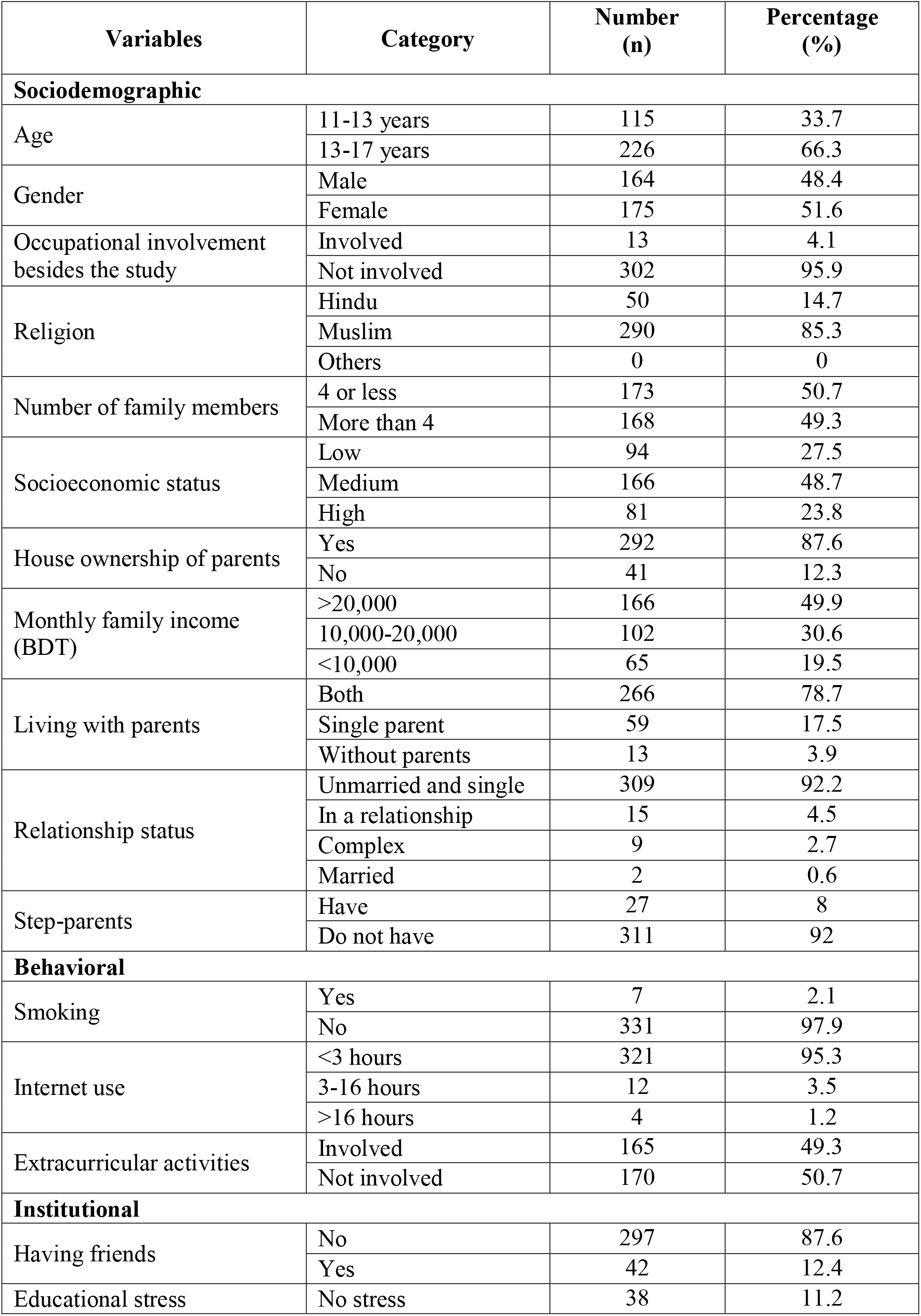
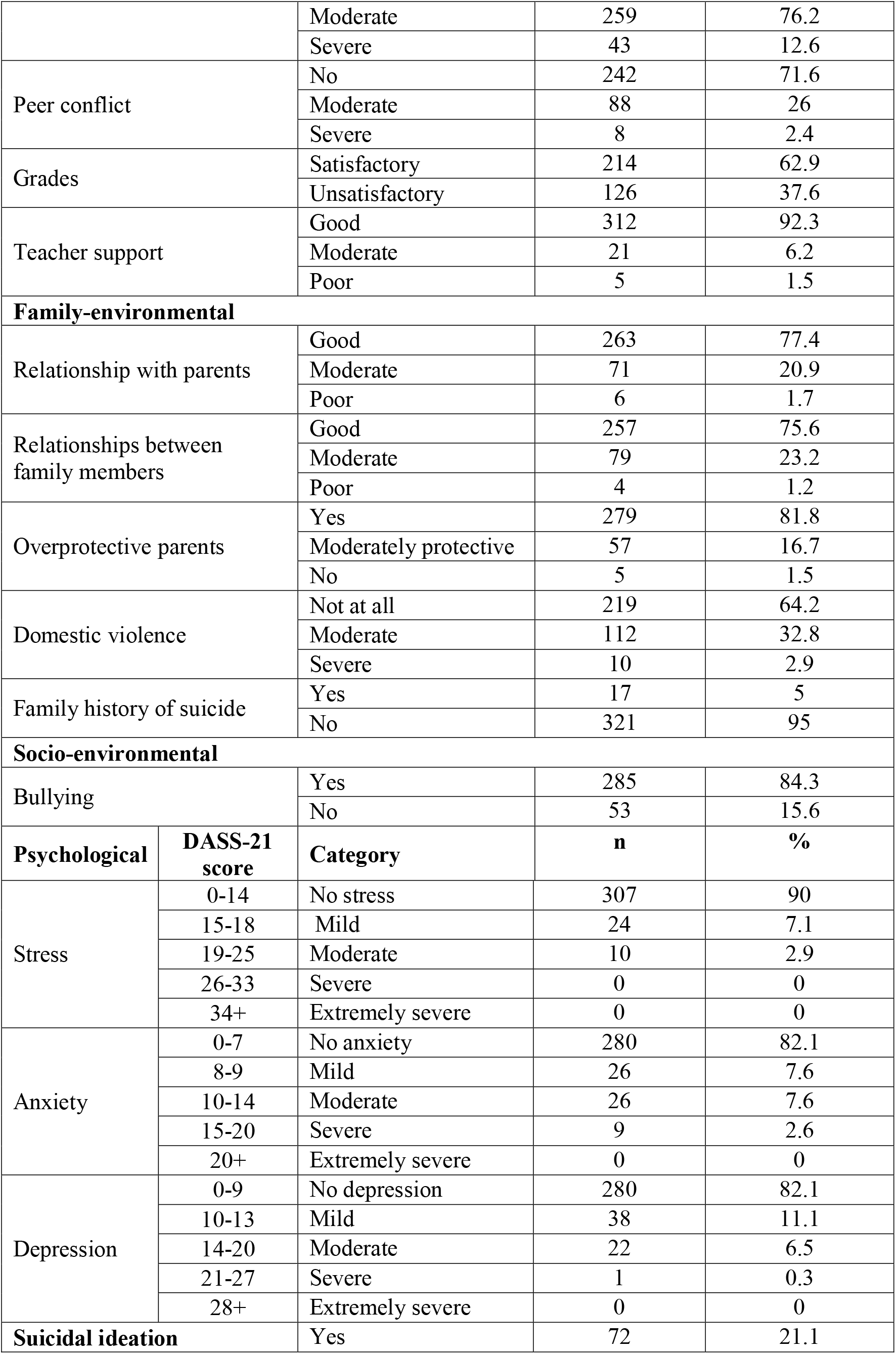
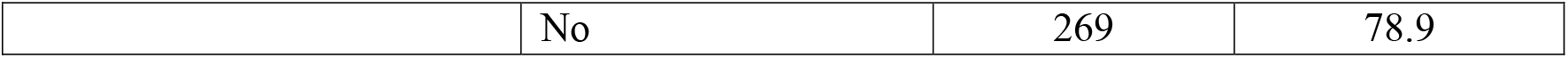
Characteristics of 11 to 17-year-old adolescents in Lohagara (a rural subdistrict in Bangladesh), 2024.

| Variables | Category | Number (n) | Percentage (%) |
| --- | --- | --- | --- |
| **Sociodemographic** |  |  |  |
| Age | 11-13 years | 115 | 33.7 |
|  | 13-17 years | 226 | 66.3 |
| Gender | Male | 164 | 48.4 |
|  | Female | 175 | 51.6 |
| Occupational involvement besides the study | Involved | 13 | 4.1 |
|  | Not involved | 302 | 95.9 |
| Religion | Hindu | 50 | 14.7 |
|  | Muslim | 290 | 85.3 |
|  | Others | 0 | 0 |
| Number of family members | 4 or less | 173 | 50.7 |
|  | More than 4 | 168 | 49.3 |
| Socioeconomic status | Low | 94 | 27.5 |
|  | Medium | 166 | 48.7 |
|  | High | 81 | 23.8 |
| House ownership of parents | Yes | 292 | 87.6 |
|  | No | 41 | 12.3 |
| Monthly family income (BDT) | >20,000 | 166 | 49.9 |
|  | 10,000-20,000 | 102 | 30.6 |
|  | <10,000 | 65 | 19.5 |
| Living with parents | Both | 266 | 78.7 |
|  | Single parent | 59 | 17.5 |
|  | Without parents | 13 | 3.9 |
| Relationship status | Unmarried and single | 309 | 92.2 |
|  | In a relationship | 15 | 4.5 |
|  | Complex | 9 | 2.7 |
|  | Married | 2 | 0.6 |
| Step-parents | Have | 27 | 8 |
|  | Do not have | 311 | 92 |
| **Behavioral** |  |  |  |
| Smoking | Yes | 7 | 2.1 |
|  | No | 331 | 97.9 |
| Internet use | <3 hours | 321 | 95.3 |
|  | 3-16 hours | 12 | 3.5 |
|  | >16 hours | 4 | 1.2 |
| Extracurricular activities | Involved | 165 | 49.3 |
|  | Not involved | 170 | 50.7 |
| **Institutional** |  |  |  |
| Having friends | No | 297 | 87.6 |
|  | Yes | 42 | 12.4 |
| Educational stress | No stress | 38 | 11.2 |

|  |  |  |  |  |
| --- | --- | --- | --- | --- |
|  |  | Moderate | 259 | 76.2 |
|  |  | Severe | 43 | 12.6 |
| Peer conflict |  | No | 242 | 71.6 |
|  |  | Moderate | 88 | 26 |
|  |  | Severe | 8 | 2.4 |
| Grades |  | Satisfactory | 214 | 62.9 |
|  |  | Unsatisfactory | 126 | 37.6 |
| Teacher support |  | Good | 312 | 92.3 |
|  |  | Moderate | 21 | 6.2 |
|  |  | Poor | 5 | 1.5 |
| Family-environmental |  |  |  |  |
| Relationship with parents |  | Good | 263 | 77.4 |
|  |  | Moderate | 71 | 20.9 |
|  |  | Poor | 6 | 1.7 |
| Relationships between family members |  | Good | 257 | 75.6 |
|  |  | Moderate | 79 | 23.2 |
|  |  | Poor | 4 | 1.2 |
| Overprotective parents |  | Yes | 279 | 81.8 |
|  |  | Moderately protective | 57 | 16.7 |
|  |  | No | 5 | 1.5 |
| Domestic violence |  | Not at all | 219 | 64.2 |
|  |  | Moderate | 112 | 32.8 |
|  |  | Severe | 10 | 2.9 |
| Family history of suicide |  | Yes | 17 | 5 |
|  |  | No | 321 | 95 |
| Socio-environmental |  |  |  |  |
| Bullying |  | Yes | 285 | 84.3 |
|  |  | No | 53 | 15.6 |
| Psychological | DASS-21 score | Category | n | % |
| Stress | 0-14 | No stress | 307 | 90 |
|  | 15-18 | Mild | 24 | 7.1 |
|  | 19-25 | Moderate | 10 | 2.9 |
|  | 26-33 | Severe | 0 | 0 |
|  | 34+ | Extremely severe | 0 | 0 |
| Anxiety | 0-7 | No anxiety | 280 | 82.1 |
|  | 8-9 | Mild | 26 | 7.6 |
|  | 10-14 | Moderate | 26 | 7.6 |
|  | 15-20 | Severe | 9 | 2.6 |
|  | 20+ | Extremely severe | 0 | 0 |
| Depression | 0-9 | No depression | 280 | 82.1 |
|  | 10-13 | Mild | 38 | 11.1 |
|  | 14-20 | Moderate | 22 | 6.5 |
|  | 21-27 | Severe | 1 | 0.3 |
|  | 28+ | Extremely severe | 0 | 0 |
| Suicidal ideation |  | Yes | 72 | 21.1 |
|  | No | 269 | 78.9 |

### Behavioral factors of adolescents

2% (7) of adolescents were smokers. 95% (321) used the internet not more than three hours a day. 3.6% (12) used moderately and stayed connected to the internet for 3-16 hours, whereas, 1.2% (4), were regarded as heavy users, who utilized the internet over sixteen hours. Almost half (49%) of students were involved in regular extra-curricular activities **Table *1***.

### Psychological characteristics of adolescents

The Depression Anxiety Stress Scale-21 (DASS-21) was used to assess the level of anxiety, depression, and stress of participants. According to the obtained score, 90% (327) of adolescents weren’t dominated by stress. The percentages of mild and moderate stress among pre-adults were 7% (24) and 2.9% (10). There was no one found to have severe or extremely severe stress. The rates of mild, moderate, and severe anxiety were 7.6% (26), 7.6% (26), and 2.6% (9) respectively. Meanwhile, 82% (280) had minimal or no anxiety. The percentage of students free from depression was also 82% (280). Among the rest, 11% (38) were enduring mild, 6.5% (22) moderate, and 0.3% (1) severe depression. **Table *1***.

### Institutional factors of adolescents

11.1% of students reported that they didn’t bear any academic stress, while moderate educational stress clutched a large portion (76.2%). However, 12.7% (43) experienced it vigorously. Among the participants, 62.9% (214) were complacent with their academic grades, whereas 37.1% (126) were not. The academic atmosphere appeared supportive to 92.3% (312) of students, meanwhile, 6.2% reported that they received moderate encouragement from their teachers, and 1.2% (5) perceived the institutional environment as absolutely unsupportive. 12.4% (42) of students were undergoing a dearth of close friendships. No peer conflict was experienced by 71.6% (242) of young adults at school. In contrast, 26% (88) encountered moderate, and 2.4% (8) grappled with severe disputes. **Table *1***.

### Family-environmental factors of adolescents

Slightly more than three-fourths (77.4%) of adolescents maintained good relationships with their parents. However, 20.9% (71) had moderate and 1.76% (6) endured strained parental relationships. 75.6% (257), 23.2% (79), and 1.2% (4) reported observing good, moderate, and extreme relationships between their family members respectively. 81.8% (279) of students stated having overprotective parents, and 16.7% (57) found their parents moderately concerning. Moreover, 1.5% (5) of students reported that their parents were perfunctory to their studies and homework. More than half (64.2 %) of the students didn’t experience any sort of domestic violence. On the contrary, 32.8% (112) of adolescents provided the information of being beaten by guardians sporadically, and 2.9% (10) experienced it habitually. However, 5% (17) of participants had family histories of committing or attempting suicide **Table *1***.

### Socio-environmental factors of adolescents

15.7% (53) of students were victims of bullying by classmates, teachers, family members, relatives, or neighbors **Table *1***.

### Prevalence of suicidal ideation among adolescents

The first question of the Suicidal Behavior-Revised Questionnaire (SBQ-R) was used to evaluate the rate of lifetime suicidal ideation among adolescents. The mean SBQ-R score was 4.7 (95% CI: 4.3-5). 21% (95% CI: 17% to 25.8%) (72) of students were found to have lifetime suicidal ideation after analysis. **Table *1***.

### Associated factors of suicidal ideation among adolescents

Sociodemographic, behavioral, psychological, institutional, family-environmental, and socioenvironmental factors were used to identify the associated factors of suicidal ideation among adolescents. Age, living with parents, relationship status, step-parents, smoking, close friends, peer conflict, peer isolation, educational stress, grades, bullying, the relationship with parents, the relationship between family members, overprotective parents, family history of suicide, stress, anxiety, and depression were found statistically significant with a p-value of <0.05 in chisquare test. These variables were further analyzed through a multivariate analysis using multinomial logistic regression to adjust the confounding factors.

Relationship status, smoking, having close friends, unfavorable family environment, and mild to moderate depression were found to be significantly associated with the development of suicidal ideation in preadults. Unmarried and single relationship status was negatively associated with this thought. However, it was the highest among married adolescents (45.5%). In contrast, 75% (9) of adolescents, who were in a relationship and 25% (2), who were overburdened by a complex relationship conveyed suicidal attitudes. Smoking also acted as a conducive factor for the development of suicidal ideation. 71.4% of students who smoked regularly, generated a lifetime suicidal thought. Having close friends played a pivotal role in deterring suicidal ideation in young adults. Meanwhile, 42.9% of participants, who were feeling a desideratum of close friendships grew suicidal thoughts. On the other hand, 44% of adolescents, enduring an unfavorable family environment because of the poor relationships between family members developed suicidal ideation. Depression was revealed as a significantly associated factor and students experiencing depression were prone to develop suicidal thoughts. The percentage of adolescents having suicidal ideation who were suffering from mild and moderate depression was 44.7% and 76.2% respectively. ***Table 2***.

**Table 2.**
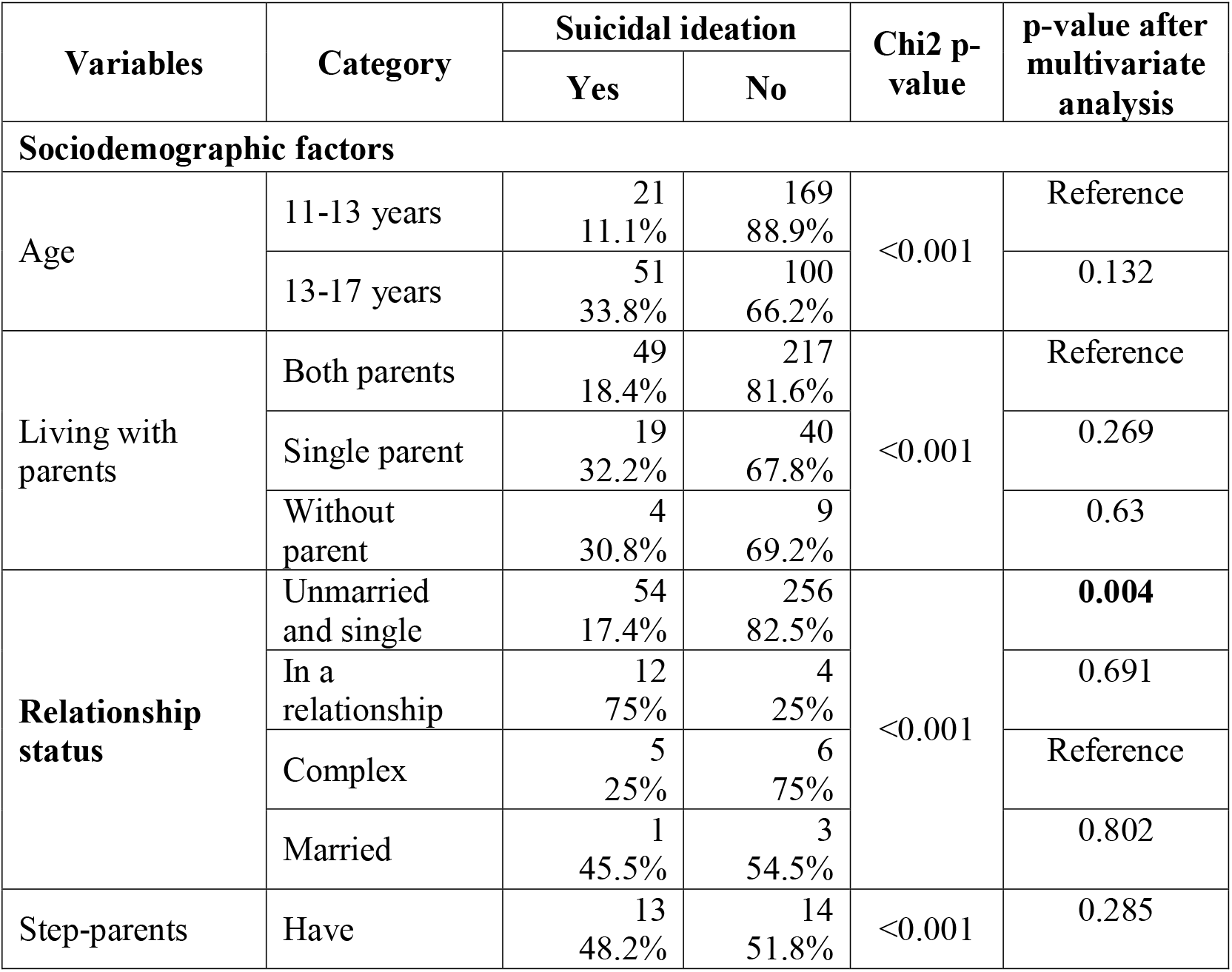
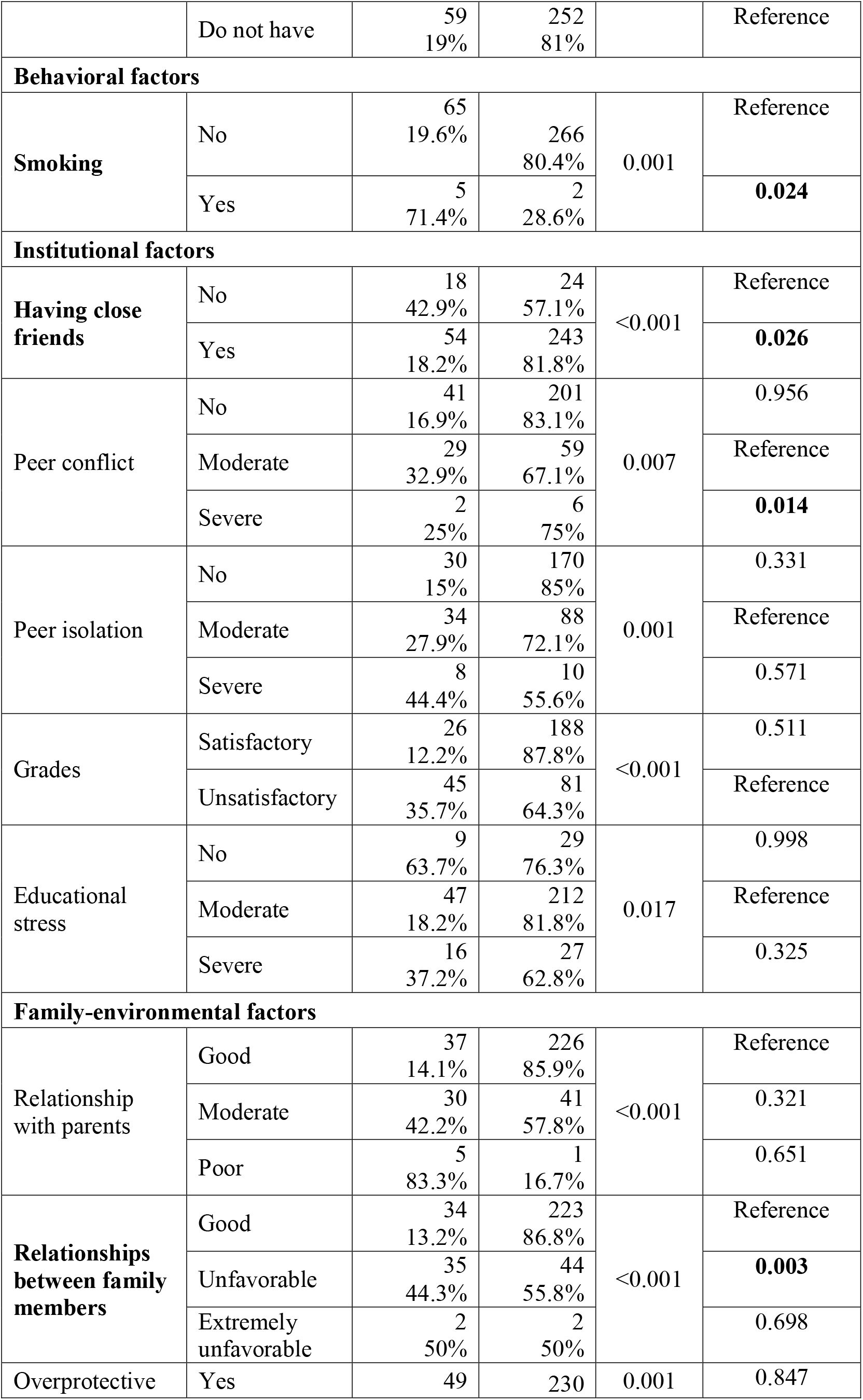
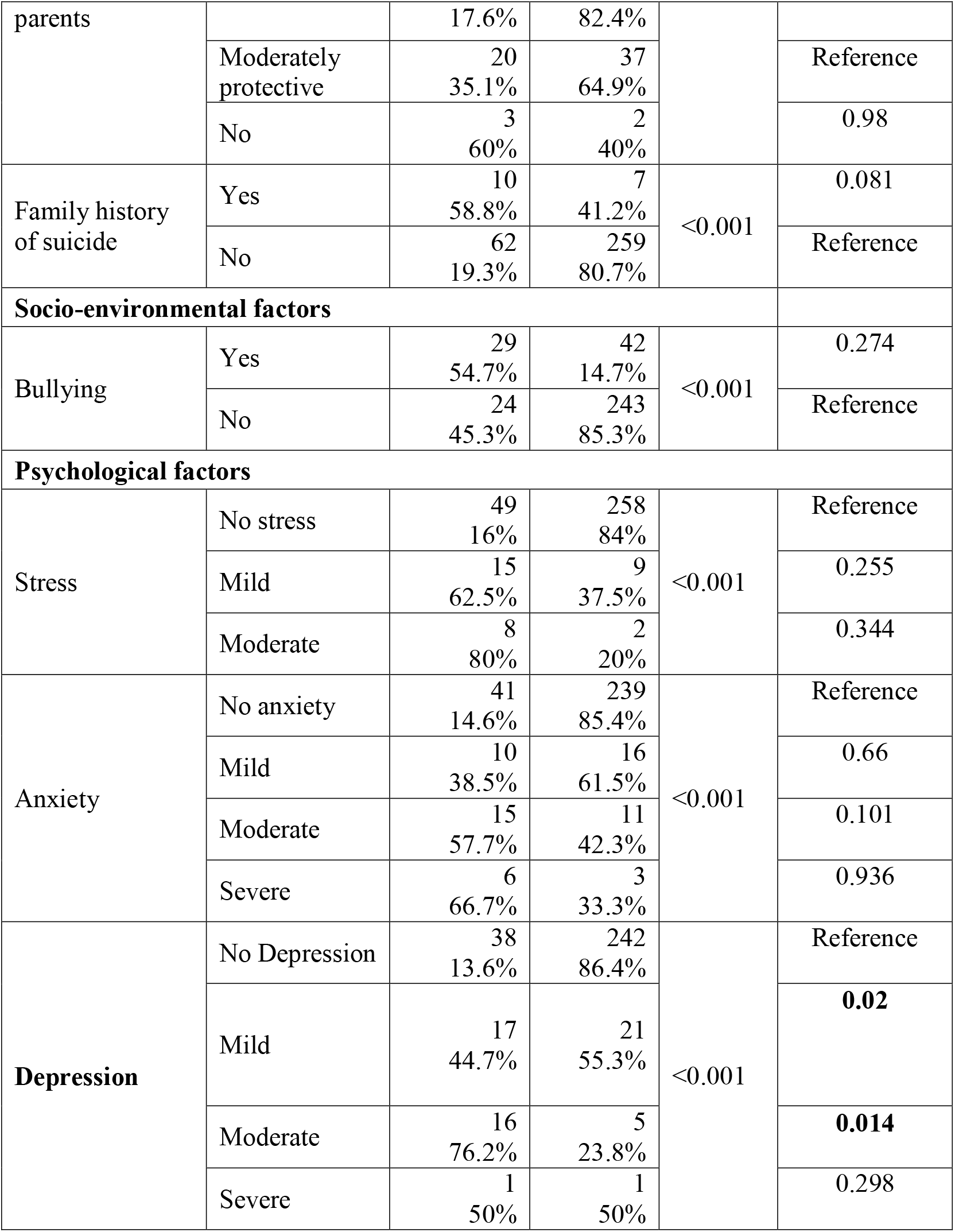
Factors associated with suicidal ideation among students of 11 to 17 years in Lohagara (a rural subdistrict in Bangladesh), 2024 (bivariate analysis by chi-squared test and multivariate analysis by multiple logistic regression).

## Discussion

This study aimed to assess the prevalence and associated factors of developing suicidal ideation among adolescents in rural Bangladesh. The point prevalence of lifetime suicidal ideation in rural adolescents is detected as 21% (95% CI: 17% to 25.8%). This percentage is attributed to a significant sociodemographic factor like relationship status. Smoking is an important behavioral factor associated with the development of suicidal thoughts in school-going adolescents. Depression, a noteworthy psychological factor is also conducive to the generation of this in 11 to 17-year-olds. In addition, an unfavorable family environment works as a crucial family-environmental factor in suicidal thought generation in this age bracket. On the other hand, having close friends, an institutional factor plays a pivotal role in nullifying the suicidal ideation of rural adolescents.

The obtained 21% prevalence from this study aligns with the 21.6% prevalence in school-going adolescents in rural Uganda (5), 20% prevalence among Korean adolescents (11), and 20.1% (95% CI: 12.6-31.7) in rural Bangladeshi adolescents (6), whereas somewhat greater than the global prevalence (14%) of suicidal ideation among 12 to 17-year-olds (3) and the prevalence in Brazilian adolescents (14.1%) (4). The probable cause might be the difference in methodology, location, and sample sizes. The evaluated result from this research is also higher than the research conducted by Khan et. al unveiling an 11.7% prevalence of suicidal behavior among 11 to 18-year-old adolescents in Bangladesh (10) and the probable cause can be the antecedent research evaluated the overall suicidal behavior, while the current one is targeted to find out the prevalence of suicidal ideation only. However, the result of this study is distant from the study conducted by Begum et. al, which revealed 5% of suicidal ideation among 14 to 19-year-olds in rural Bangladesh (7), and that is probably due to the variation in time and age groups.

Married, promised-to-get-married, or those who have received marriage requests are more prone to generate suicidal thoughts than adolescent girls who haven’t undergone the marriage process (12). This research depicts a significant association between relationship status and suicidal ideation in young adults; unmarried and single students are less likely to develop suicidal ideation at a young age.

Smoking enhances the chance of developing suicidal thoughts among preadults (13). In a study, Slomp et al. stated that female smokers have more likelihood of generating suicidal ideation (14). This study also states smoking is a contributing factor to developing this.

Lack of peer support has a decisive role in preadult suicidal thoughts development (10, 15). In a study, Biswas et. al highlighted the scarcity of close friends as a reinforcing factor for the development of suicidal behavior in adolescents (3). This study found a notable association between lack of close friendship and the generation of suicidal ideation in preadults.

Depression is revealed as a substantial psychological factor contributing to the growth of suicidal thoughts in young adults in many studies (4, 5, 16) and this study is in agreement with this. According to the current study, mild to moderate depression works as a crucial stimulator in suicidal ideation generation in adolescents.

Suicidal ideation can be developed due to a hostile family environment that is created by poor relationships between family members. Like this study, some other studies also showed an adverse family environment played a stimulating role in generating suicidal ideation in preadults(17, 18).

Some studies found significant associations between loneliness (19), bullying (20) anxiety(9, 10) stress(9), overprotective parents(21, 22), family history of suicide(23), and gender (5) with the development of suicidal tendencies in adolescents. This study doesn’t find any considerable association with them, perhaps because of the distinction in age groups, locations, and sample sizes.

## Limitation

This study was conducted with data collected from a specific school in an area because of the time and resource limitations. Though the school admission policy allows diversity, students from remote areas of Lohagara aren’t inclined to get enrolled here. For this reason, this study doesn’t fully capture the divergence of the community people and thus leaves a chance of generalizability bias.

## Conclusion

However, about 2 in 10 students are developing suicidal ideation at an early age, which is a matter of concern. Further research with students from different locations in Bangladesh will help identify other associated or area-specific factors if available. Stakeholders and policy-makers should come forward to draft plans and policies to scale down the burden. The attitude and behavior of adolescents should be monitored carefully by their parents and teachers. Early screening and interventions are fundamental to protect them from making a dreadful decision. In addition, ensuring the participation of young adults in health promotional and health educational programs organized on this issue and increasing their creative opportunities may keep their mental health well and safeguard them from emerging suicidal tendencies at school age.

## Data Availability

data can be found in figshare
DOI: 10.6084/m9.figshare.25854445

https://figshare.com/articles/dataset/Prevalence_and_associated_factors_of_suicidal_behavior_of_adolescents_in_rural_Bangladesh/25854445

## Acknowledgment

I am profoundly thankful to the Almighty for granting me the opportunity to pursue an MPH (Epidemiology) at North South University. I am also grateful to North South University for allowing me to do the research. I am greatly thankful to S M Hayatuzzaman, Headmaster, Lohagara Government Pilot High School Lohagara, Narail for his unwavering support to carry out the study.

